# Evaluation of System One Models on Biomedical Texts

**DOI:** 10.64898/2026.11.15.26368126

**Authors:** Anthony F. Chen, Albert Yang, Daniel R. Guth, Frederick M. Berl, Stephen C. Guth, William Yuan

## Abstract

**Objective:** To compare the performance and cost of System One models (SOMs) with general-purpose large language models (LLMs) on biomedical tasks and evaluate confidence-based routing.

**Materials and Methods:** We evaluated three SOMs and three LLMs across seven datasets and eight endpoints covering question answering, error detection, structured extraction, and clinical-note assessment. Outcomes included task-specific performance, recorded inference cost and latency, and confidence calibration. Calibration data determined routing schemes and confidence thresholds to evaluate test predictions, with cost-matched random referral as a comparator.

**Results:** Claude Opus 5.5 had the highest primary performance on all eight endpoints. The strongest SOM, Jev, had competitive performance with the evaluated LLMs and cost less than 1% of Opus on most datasets. Three selected SOM-to-LLM routes reduced costs by 30.0%-71.7% relative to the receiving LLM, with modest reductions in performance. On one benchmark, SOM-to-SOM routing achieved 91.1% accuracy, compared with 82.3% and 82.2% for the individual models.

**Discussion:** SOMs offered favorable performance-cost tradeoffs for most tasks. The magnitude of the advantages depended on task structure and prompting requirements. Confidence-based routing between SOMs and LLMs can leverage complementary model strengths and offers additional flexibility in performance-cost tradeoffs. The price advantages offered by SOMs can facilitate experimentation and development of workflows. This warrants further evaluation on closed datasets.

**Conclusion:** For biomedical tasks with predefined choices, SOMs represent a competitive stand-alone or first-stage approach.

## BACKGROUND AND SIGNIFICANCE

Large language models (LLMs) have shown promise across many biomedical applications,[1–4] but their freeform text outputs may not be optimal for the many tasks that require only a categorical decision or numerical estimate. Better approaches to these simpler tasks may exist, especially given the cost, latency, and reliability considerations of LLM-generated general-purpose text.[5,6]

A recent development in cognitive artificial intelligence architecture is “System One” decision models (SOMs). These are named after “System 1 thinking”: the human brain’s fast, intuitive information processing system, built on heuristics and biases.[7,8] SOMs accept text along with a question and predefined set of answers, and then return probabilities over those answers. Although SOMs can still select an incorrect answer, the predefined response space limits the types of possible error that can be made. This makes it easier to characterize those errors and their consequences, then compensate accordingly, similar to how clinicians utilize diagnostic tests or risk scores with known error profiles.[9–11]

Because SOMs are narrower in scope than LLMs, they are also substantially cheaper per token. Examples of new SOMs include Jev (TypeSafe AI), Clef (Cloudflare, built from Qwen3.8–27B), and Clef-flash (Cloudflare, built from Qwen3.5–9B).[12,13] To our knowledge, there is limited evidence on how SOMs perform relative to LLMs for biomedical applications, and a comparative analysis could identify use cases where SOMs may function as alternatives or supplements to LLMs.

## OBJECTIVE

In this study, we scored three SOMs and three general-purpose LLMs on established biomedical benchmarks across seven datasets, covering applications like named entity recognition, relation extraction, text classification, question answering, document summarization, and simplification/translation. Furthermore, we explored how selectively routing SOM predictions to other models could impact tradeoffs between cost and performance.

## MATERIALS AND METHODS

We evaluated six models across eight benchmarking tasks from seven datasets (two benchmarks were derived from PriMock57). The datasets used to test SOMs and LLMs are summarized in Table 1. The models evaluated are summarized in Table 2. The prompts utilized are in Supplementary Table 1.

**Table 1:** Summary of evaluation tasks.

| Dataset | Input and requested decision | Origin and language | Calibration size | Test size | Endpoint |
|---|---|---|---|---|---|
| **PubMedQA**<br>Jin et al. 2019 | Question and context; choose yes, no, or maybe. | Expert-labeled biomedical literature questions; English. | 500 questions | 499 questions | Accuracy (%) |
| **MedHallu**<br>Pandit et al. 2025 | Question, context, and candidate answer; classify if factual or hallucinated. | Paired factual and constructed hallucinated answers; English. | 1,000 judgments | 2,000 judgments | Accuracy (%) |
| **MEDEC-MS**<br>Ben Abacha et al. 2025 | Exam-derived clinical text; detect presence of a medical error. | Medical exam questions with expert-introduced errors; English. | 574 texts | 597 texts | Accuracy (%) |
| **TCGA-242**<br>Chang et al. 2026 | Surgical pathology report; extract categorical field values. | Expert-verified annotations from public cancer dataset; English. | 85 reports | 152 reports | Micro-F1 |
| **PriMock57**<br>**Incorrect statements**<br>Papadopoulos Korfiatis et al. 2022; Moramarco et al. 2022 | Transcript and candidate Clinical note; estimate number of incorrect statements. | Expert verified counts from simulated consultations; English. | 70 notes | 215 notes | Spearman ρ |
| **PriMock57**<br>**Omissions**<br>Papadopoulos Korfiatis et al. 2022; Moramarco et al. 2022 | Transcript and candidate Clinical note; estimate number of omissions. | Expert verified counts from simulated consultations; English. | 70 notes | 215 notes | Spearman ρ |
| **OmissionBench**<br>Fox et al. 2026 preprint | Transcript and clean or perturbed note; identify if an error is present. | Released clean or constructed-error pairs from simulated consultations; English. | 44 pairs | 114 pairs | Paired discrimination (%) |
| **IMCS21**<br>Chen et al. 2023 | Chinese pediatric consultation dialogue; infer the status of 331 normalized symptoms. | Expert-defined annotations; Chinese. | 833 dialogues | 811 dialogues | Labeled micro-F1 |

**Table 2:** Summary of evaluated models.

| Model | Type | Developer | Cost<br>Input / output (USD per million input/output tokens) | Release date | Notes |
|---|---|---|---|---|---|
| Jev 1.13.0 | SOM | TypeSafe AI | 0.042 / free | 2026-09-15 | Proprietary decision model |
| Clef | SOM | Cloudflare | 0.24 / free | 2026-10-01 | Post-trained from Qwen3.8-27B; open weight |
| Clef-flash | SOM | Cloudflare | 0.038 / free | 2026-10-01 | Post-trained from Qwen3.5-9B; open weight |
| Haiku 4.5 | LLM | Anthropic | 1.00 / 5.00 | 2025-10-15 | Proprietary general-purpose generative model |
| Haiku 5.5 (medium effort) | LLM | Anthropic | 0.10 / 0.50 | 2026-10-07 | Proprietary general-purpose generative model |
| Opus 5.5 (medium effort) | LLM | Anthropic | 4.00 / 20.00 | 2026-09-22 | Proprietary general-purpose generative model |

**PubMedQA** contains research questions derived from PubMed abstracts with a clinical question in the title and the answer in the conclusion.[14] Models received the title question and abstract context without the conclusion, and predicted whether the conclusion resolved to “yes,” “no,” or “maybe.” One question in the test set (PMID 16776337) was excluded because the supplied abstract context lacked the results needed to answer it.

**MedHallu** was derived from PubMedQA, and contains biomedical questions, answers, and constructed plausible hallucinations across the following categories: misinterpretation of question, incomplete information, mechanism and pathway misattribution, and methodological and evidence fabrication.[15] Models were given the question and context followed by either the correct answer or a hallucination, then predicted which they had received.

**MEDEC-MS** contains medical examination-derived clinical texts which were either entirely correct or contained a single author-injected medical error involving diagnosis, management, treatment, pharmacotherapy, or causal organism.[16] Models would receive the text and attempt to predict the presence or absence of an error.

**TCGA-242** contains surgical pathology reports and structured annotations from The Cancer Genome Atlas.[17] Models provided with the reports aimed to identify whether the report described a primary cancer excision and if so, what organ was involved, then extracted a set of organ-specific fields based on the selected organ. Expert review of the labels and notes identified inconsistencies regarding the availability of the surgical approach field from some of the reports, which were corrected. Furthermore, five records were duplicated, which we removed. Our corrected version of TCGA-242 is provided in the Supplement.

**PriMock57** is a dataset comprised of 57 simulated primary care consultations, using case cards describing symptoms with a clinical story.[18] A panel of five clinicians graded 285 generated notes describing the consultations and identified contradictions, hallucinations, repetitions, and omissions within the notes.[19] Models received the initial simulated consultation transcript and a candidate note, and estimated the number of incorrect statements or omitted items. These two tasks were evaluated separately.

**OmissionBench** is a clinical-documentation error benchmark derived from Primock57. It contains transcripts paired with two versions of notes: one unchanged, and one containing an introduced error. Models assessed each transcript and note-version separately, then were evaluated on whether their identified error probability for the erroneous note was higher than for the unchanged note.[20]

**IMCS-21** is a Chinese-language medical dialogue dataset that consists of pediatric consultation dialogues over 10 diseases.[21] We used the symptom label detection task, which identifies symptoms mentioned in the dialogues and labels them as present or absent in the patient, or not mentioned at all in the dialogue. The dataset contained an inventory of 331 symptoms. For SOMs, the task was administered through 331 questions, one for each symptom. Clef and Clef-flash required 6 separate requests, each repeating the dialogue and context, to complete all 331 questions due to API limitations. LLMs received the entire symptom inventory as part of a single prompt.

For each task, questions were formatted into context and answer choices. SOMs were prompted with these directly, and model choices and confidence values were evaluated directly. LLMs received the context and answer choices, formatted into a single prompt, with the addition of a request to generate a confidence estimate. We preserved the developer-defined test splits when provided. Otherwise, we constructed calibration/test partitions, keeping reports from the same source and notes from the same consultation within a single partition.

Primary evaluation metrics included cost, time, calibration, and performance metrics specific to each benchmark. For LLMs, a prediction was considered a failure when no valid response was generated, including refusals, truncated or malformed outputs, and responses that did not satisfy the required answer schema. For classification tasks, failures were scored as incorrect. For TCGA-242 and IMCS21, failures resulted in no extracted values. For OmissionBench, a pair received zero discrimination credit if either note’s prediction failed. For PriMock57, failed predictions were assigned a count of zero for Spearman correlation calculation.

Based on performance in the calibration set, we further evaluated tandem approaches, where a SOM is first applied to every question, then low confidence answers are routed to an LLM on the performance/cost Pareto frontier. Questions that resulted in a confidence level below a certain threshold (the SOM referral threshold, q) had their answers compared to the same question that had been provided to an LLM. The LLM prompts did not mention that the questions had been routed from an SOM. The routing model pairs and SOM referral thresholds were evaluated in the calibration set, and schemes that offered improvement were evaluated in the test set.

## RESULTS

### SOMs achieve strong predictive performance relative to inference cost

Opus 5.5 achieved the strongest primary performance across all eight tasks but also had the highest inference costs. Among SOMs, Jev achieved the highest or tied-highest point performance on seven endpoints, while Clef performed best on TCGA-242 (Figure 1, Supplementary Table 2). Jev approached the performance of Opus most closely on PriMock57, with Spearman correlations of 0.809 versus 0.825 for omissions and correlations of 0.828 versus 0.881 for incorrect statements. On TCGA-242, Jev scored a micro-F1 of 0.790 versus 0.839 for Opus. Detailed results are presented in the Supplement (Supplementary Tables 2-12).

**Figure 1:**
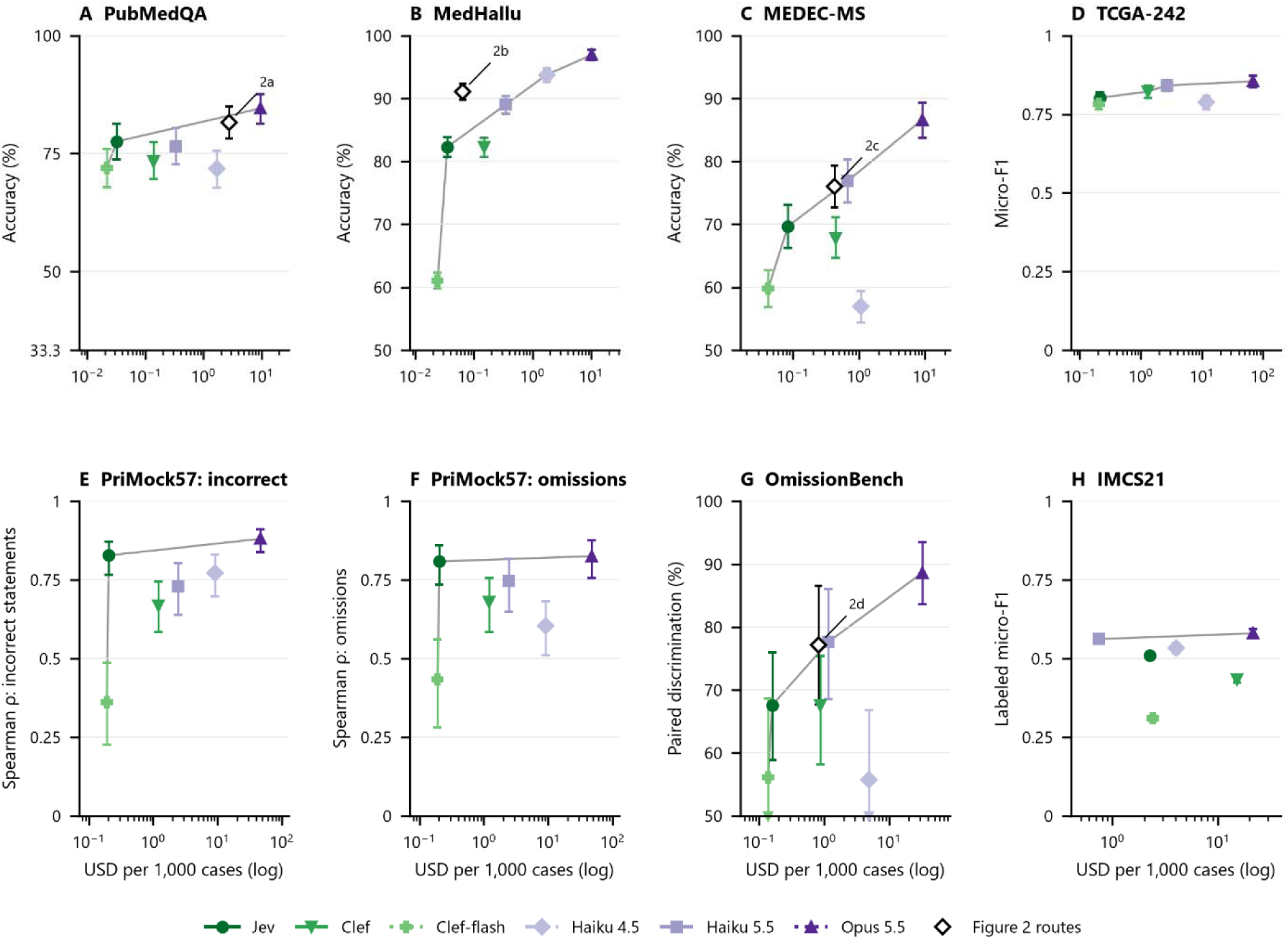
Comparison of test set performance versus cost between SOMs (green) and LLMs (purple). The Y-axes are scaled for each benchmark, with the lower limit the expected performance if a model is randomly guessing from among the answer choices and the upper limit equal to complete accuracy on the task. Error bars represent 95% CIs. Gray lines represent the Pareto frontier. Alt text: Eight graphs demonstrate the performance versus cost of the tested models, for each benchmark. LLMs cost much more but perform slightly better.

Over all tasks, calibration varied substantially by task and model (Supplementary Tables 3A and 3B), but SOMs had substantial latency advantages (Supplementary Table 5). Opus 5.5 performed tasks in 5.11-25.11 seconds, while Jev took only 0.16-0.34 seconds, aside from the IMCS21 benchmark. This corresponded to a 25-90x increase in speed, depending on task.

Except for IMCS21, Jev’s inference cost ranged from 100 to 320 times lower than Opus 5.5 and 7 to 13 times lower than Haiku 5.5. The specific performance and cost differences varied substantially by task (Supplementary Table 4).

With IMCS21, Haiku 5.5 achieved higher labeled micro-F1 than Jev (0.562 versus 0.510) at lower cost ($0.746 versus $2.260 per 1,000 dialogues) and was cheaper than all three SOMs. The reduced cost advantage of SOMs with IMCS21 may reflect its large symptom inventory, as evaluating 331 symptom labels required substantially more prompting than the other tasks. Due to the nature of needing to prompt regarding every symptom, this necessarily resulted in an inefficient evaluation harness. However, there were high LLM failure rates as well: Haiku 4.5 failed in 11% of cases, while Haiku 5.5 failed in 1.3% (Supplementary Table 12). All evaluated LLMs and SOMs (recognition F1 from 0.420-0.758) also performed below the BERT-based models reported in the original benchmark (recognition F1 = 0.916).[17,21]

### Tandem systems with confidence-based routing improve cost-performance trade-offs

We evaluated routing on four out of eight benchmark tasks, focusing experiments on tasks where either i) the calibration performance-cost Pareto frontier suggested an opportunity to improve performance of a low-cost SOM by selectively escalating cases to an LLM, or ii) we observed that two different SOMs had substantially different error profiles in the calibration data. We did not evaluate routing on TCGA-242 or the two PriMock57 endpoints because Pareto-efficient SOMs already approached LLM performance, leaving less opportunity for improvement through escalation. Additionally, IMCS21 was excluded because no SOM lay on the Pareto frontier. For all other tasks, the routing schemes found to be Pareto-efficient in calibration data improved test performance compared to the SOM alone (Figure 1; Table 3). All learned routing schemes were also found to be Pareto-efficient in the test set (Supplementary Figure 1). We summarize four selected routing schemes in Figure 2, and detailed results are presented in the Supplement (Supplementary Table 13).

**Figure 2:**
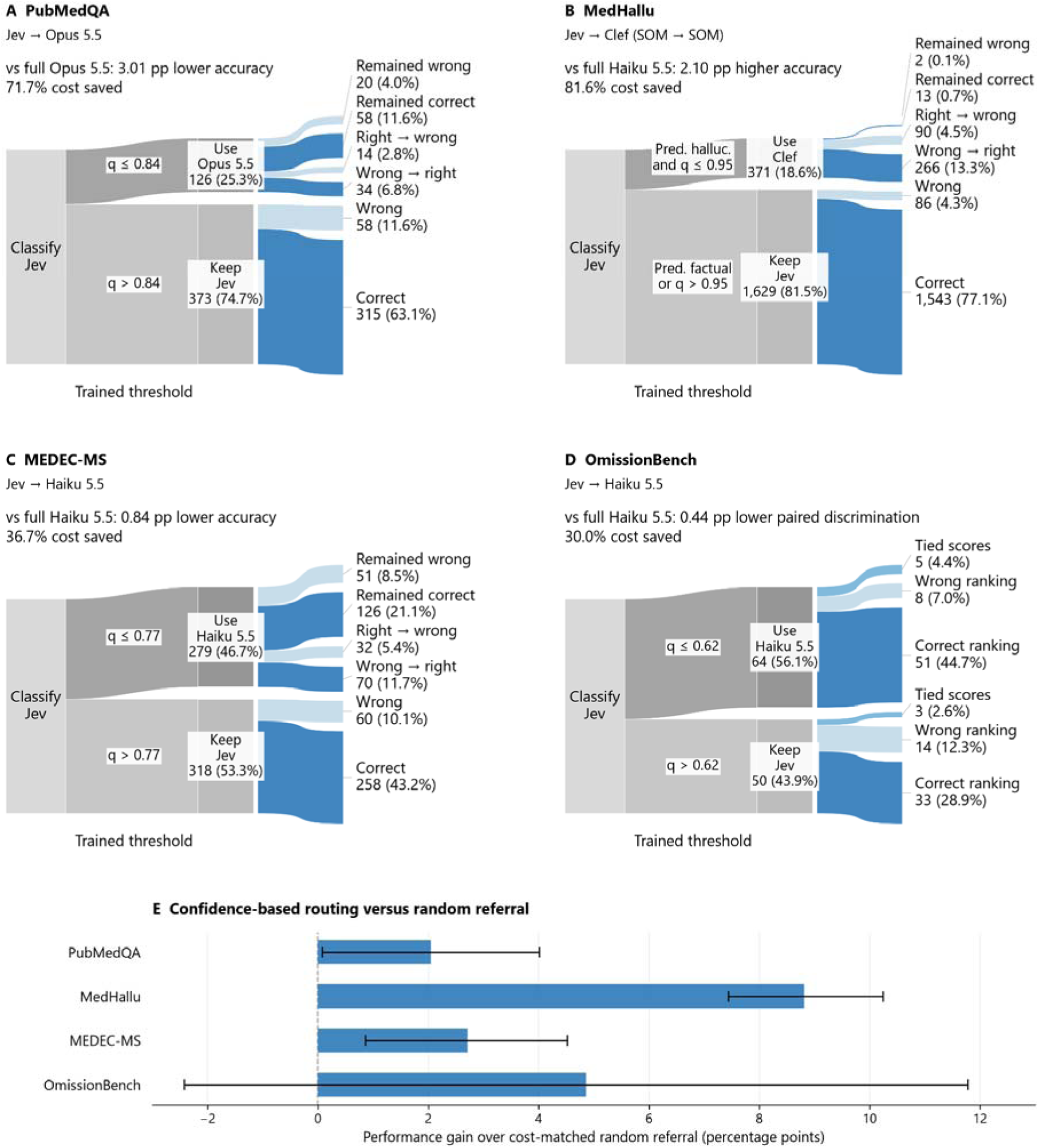
Routing performance of SOMs and LLMs. Use of SOM confidence estimates to determine escalation to LLM leads to stronger performance at lower cost. Cutoffs were determined from calibration data. Alt text: Four graphs depict the breakdown of tasks that were routed by a SOM to an LLM, with a representation of how many were changed, and which ended up being correct.

**Table 3:** Summary of selected routing experiments.

| Dataset | Routing scheme | Cal perf. (%) | SOM referral threshold | Test perf. % (95% CI) | Test cost USD/1,000 | Test referred (%) | Performance vs. nearest LLM | Cost saved vs. nearest LLM |
|---|---|---|---|---|---|---|---|---|
| PubMedQA | Jev → Opus 5.5 | 83.40 | q ≤ 0.84 | 81.56 (78.16–84.97) | 2.7525 | 25.3 | Opus 5.5 -3.01 pp | 71.7% |
| MedHallu | Jev → Clef Hallucination-only | 91.60 | q ≤ 0.95 | 91.10 (89.85–92.30) | 0.0636 | 18.6 | Haiku 5.5 +2.10 pp | 81.6% |
| MEDEC-MS | Jev → Haiku 5.5 | 77.00 | q ≤ 0.77 | 76.05 (72.73–79.37) | 0.4269 | 46.7 | Haiku 5.5 -0.84 pp | 36.7% |
| OmissionBench | Jev → Haiku 5.5 | 87.50 | q ≤ 0.62 | 77.19 (67.70–86.54) | 0.8183 | 56.1 | Haiku 5.5 -0.44 pp | 30.0% |

On PubMedQA (Figure 2A), routing 25.3% of questions from Jev to Opus 5.5 recovered roughly half of the performance differential between Jev and Opus, while maintaining a 71.7% lower cost than exclusively using Opus. Among referred questions, Opus corrected 34 Jev errors and introduced 14 errors, yielding a net gain of 20 correct answers. On MEDEC-MS and OmissionBench (Figure 2C,D), routing from Jev to Haiku 5.5 recovered nearly the entire performance differential between the two models. On MedHallu, in the calibration set, Jev showed high sensitivity for hallucinations (95.0%) but lower specificity for factual answers (68.4%), whereas Clef showed lower sensitivity (64.6%) and near-perfect specificity (99.8%). A routing scheme where low-confidence Jev hallucination predictions were routed to Clef improved accuracy by nearly 10 percentage points (Jev: 81.7%; Clef: 82.2% → 91.6%). The joint SOM outperformed Haiku 5.5 by 2.1 percentage points at less than 20% of the cost (Table 3; Figure 2B). We further identified that SOM confidence-based routing outperformed cost-matched random referral (Figure 2E) for 3 out of 4 tasks, supporting the value of selecting which cases to route. Finally, for all routing schemes besides OmissionBench, the first-line SOM consistently referred “harder” cases to the second-line model. For example, on PubMedQA, for questions where Jev’s answer was retained, Jev scored 84.5% compared to Opus’s 88.5%. However, Opus only scored 73% on the questions routed from Jev. (Supplementary Table 14).

## DISCUSSION

Across eight biomedical benchmarks, SOMs offered substantial reductions in inference cost and latency, with competitive predictive performance relative to the evaluated LLMs. Opus 5.5 achieved the highest primary performance with every endpoint, while Jev was the strongest SOM on seven of eight endpoints. Selective routing between SOMs and LLMs recovered part of these performance differences while retaining cost savings. The value of these advantages depends on how the output will be used. A model that extracts structured data from radiology reports may have different performance requirements from one that directly guides a clinical action. These findings support choosing models based on the workflow requirements of a specific task rather than performance alone, including error handling processes, cost, volume, and response format.

SOMs may be particularly useful for tasks that can be expressed as bounded decisions over predefined choices. Examples we evaluated include classifying texts, identifying errors, and extracting information from unstructured data. Their low inference costs could make repeated assessment of large document collections more feasible, while their short recorded latencies could support interactive or multistep applications.

For datasets with larger SOM–LLM performance gaps, confidence-based routing improved performance of the SOM without needing a larger, costlier LLM for every case. We also observed substantially different error profiles among SOMs, creating opportunities to combine models with complementary strengths. The low cost of SOM inference makes these multistep workflows feasible within budgets that might otherwise permit only a single LLM assessment. Beyond routing, this could allow repeated assessments with different prompts or decomposition of a task into several bounded decisions. Lower costs also make it more affordable to compare prompting strategies and workflow designs during development. Although we evaluated only selected routing schemes, our findings suggest that the value of SOMs extends beyond just inexpensive individual predictions, such as enabling more flexible use of a fixed model budget.

The IMCS21 results show how the formulation of a task can limit the practical advantages of SOMs. Our implementation required SOMs to assess all 331 symptoms for every dialogue, without first narrowing the candidate set using the consultation context. Performance on this task may not be representative of workflows where relevant questions are specified in advance or candidate symptoms can first be filtered. The resulting query burden offset the inherent low per-token cost of SOMs, and Haiku 5.5 achieved higher labeled micro-F1 at lower cost than all three SOMs. Furthermore, all evaluated SOMs and LLMs underperformed the BERT-based benchmarks. Interpretation of performance is also complicated by overlapping symptom categories. Clinician inspection identified broad and specific concepts, such as “fever” and “moderate fever,” that were not consistently nested in the reference annotations. Exact-match scoring may therefore penalize clinically compatible predictions. A clear understanding of the intended workflow, use case, and error mitigation strategies for a model is crucial to evaluating performance and identifying optimal tasks that SOMs, LLMs, or tandem systems are well-suited for.

Our study has several important limitations. The evaluation used public benchmarks rather than prospective clinical workflows. We cannot rule out the possibility that the SOMs or LLMs we tested were originally trained on the benchmarks evaluated. Some benchmarks also shared underlying source material and so do not represent fully independent evaluations: MedHallu was derived from PubMedQA, and OmissionBench used PriMock57 consultations. Cost comparisons reflect the evaluated API-based deployments and pricing assumptions and may differ from those achievable through self-hosting. In particular, Clef and Clef-flash can be self-hosted, where costs would depend on hardware, utilization, and operational overhead rather than API prices alone. Reference annotations were also imperfect. Expert review identified inconsistencies in the availability of the surgical approach field in TCGA-242, which we corrected before evaluation. These corrections limit direct comparability with evaluations using the original labels.

## CONCLUSION

Overall, SOMs are a competitive approach for many biomedical tasks, particularly working in tandem with LLMs or other SOMs. We highlight how the choice of model for a task is dependent on the integration context, and how SOMs represent a tool with a dramatically different cost-benefit profile. These results support further evaluation of SOMs for these tasks and exploration of more complex workflow development.

## Supporting information

Corrected TCGA Dataset

Experiment Results

## Data Availability

All data produced in the study is either uploaded along with the manuscript or available upon reasonable request to the authors.

## Acknowledgments

Code was written and data was processed by the artificial intelligence models described in the Methods.

